# The main factors influencing COVID-19 spread and deaths in Mexico: A comparison between Phases I and II

**DOI:** 10.1101/2020.12.22.20248716

**Authors:** Francisco Benita, Francisco Gasca-Sanchez

## Abstract

This article investigates the geographical spread of confirmed COVID-19 cases and deaths across municipalities in Mexico. It focuses on the spread dynamics and containment of the virus between Phase I (from March 23 to May 31, 2020) and Phase II (from June 1 to August 22, 2020) of the social distancing measures. It also examines municipal-level factors associated with cumulative COVID-19 cases and deaths to understand the spatial determinants of the pandemic. The analysis of the geographic pattern of the pandemic via spatial scan statistics revealed a fast spread among municipalities. During Phase I, clusters of infections and deaths were mainly located at the country’s center, whereas in Phase II, these clusters dispersed to the rest of the country. The regression results from the zero-inflated negative binomial regression analysis suggested that income inequality, the prevalence of obesity and diabetes, and concentration of fine particulate matter (PM 2.5) are strongly positively associated with confirmed cases and deaths regardless of lockdown.

## 1. Introduction

As of November 24, 2020, Mexico ranks fourth in the world in coronavirus disease 2019 (COVID-19) deaths, behind only by the U.S., Brazil and India. The country has a total 1’049,358 positive cases and 101,926 deaths, which at present, represents one of the highest lethality rates (10.29%) in the world. Mexico, like other Latin American countries is framed by growing poverty levels, large income inequality and disparities in access to healthcare services (Dávila-Cervantes and Agudelo-Botero, 2019; Hessel et al., 2019; Mesenburg et al., 2018). Furthermore, the prevalence of hypertension, diabetes, and other conditions that might shape obesity in the country have a much larger role on the lethality disease. The strong relationship between being individual with overweight or obesity and severe outcomes of hospitalization has been well documented (Hernández-Garduño, 2020; Popkin et al., 2020). Hence, individuals with excessive body fat, or major cardiometabolic problems (ranging from hypertension to cardiovascular disease and diabetes) could present severe or even lethal complications. In this vein, Mexico faces a significant risk as, for years now, it has been in the second spot when it comes to the global ranking of adult obesity, just behind the U.S., whereas, overweight and obesity in childhood rates are the highest worldwide (OECD, 2019).

Similarly, an enormous amount of scientific literature has provided strong evidence that climate factors such as high air temperature or exposure to air pollution lead to an increase of lethality rates (Lin et al., 2020a,b; Ahmadi et al., 2020; Bashir et al., 2020; Riccò et al., 2020; Sobral et al., 2020). This has been explained by the contribution of air pollution to respiratory tract infections, pulmonary disease, or diabetes burden (Lubrano et al., 2020; Ma et al., 2020; Xu et al., 2020). Mexico, with vastly different climates among its states and municipalities, has presented differentiated effects on COVID-19 confirmed cases (Méndez-Arriaga, 2020). Some other studies have shown the underlying socio-economic conditions could correlate to the dynamics of the pandemic. For instance, Bambra et al. (2020) documented how the exacerbated existing social inequalities in the country have translated to higher rates of infection and mortality among the most disadvantaged communities. One can also hypothesize that, like other highly densely populated cities in the world (Sun et al., 2020a,b,c), the relative number of COVID-19 infections and deaths in Mexican metropolitan areas would be larger than non-metro areas.

On top of the above-mentioned socio-economic, health and climate factors associated with the dynamics of the disease, physical distancing and lockdown measures restricting mobility add an extra layer of complexity to the understanding of the unfolding of the pandemic. On one side of the spectrum, government authorities of countries like China, Germany or South Korea acted early by sending their residents a clearly message about what actions to be taken. The positive impact of lockdown on containing the COVID-19 outbreak has been observed in China (Lau et al., 2020), Italy (Signorelli, 2020), Hong Kong (Lam et al., 2020) United Kingdom (Ferguson et al., 2020) or Thailand (Dechsupa et al., 2020). On the other side of the spectrum, government’s actions in countries like the U.S., Brazil or Mexico have been questioned for failing to adopt early and strict containment. In Mexico, “Phase I” of the temporary mobility restriction efforts started with the National Safe and Social Distancing Program (in Spanish *Jornada Nacional de Sana Distancia*), and comprised 14 weeks between March 23th and May 31st, 2020. During Phase I, residents were exhorted to: (i) to stay at home (not enforced), (ii) wash their hands repeatedly, and (iii) avoid touching their face. With a 56% of informal employment, the possibility to “stay home” was limited to a small sub-group of Mexicans. In fact, reports of automobile traffic, tracking of mobile phones and social media activities indicated that most people were ignoring Phase I mobility restrictions (Macip, 2020).

This study investigates main socio-economic, health, climate and mobility factors associated to COVID-19 infections and deaths. To do so we fit Zero-Inflated Negative Binomial (ZINB) regression models. We also attempted to understand the Relative Risk (RR) of the transmission dynamics of current and future emerging geographical clusters. For this task, we implemented discrete Poisson probability models. The RR is a non-negative number representing how much more common disease is inside a geographical cluster compared to outside. We use data from the 2,459 Mexican municipalities to explore correlations of variables during Phase I and II. Here, Phase II is defined as the following 14 weeks after Phase I ended, this is, between June 1st and August 22th, 2020.

Our Space-Time Scan Statistics (SaTScan) allows the detection, size and duration of emerging clusters of cumulative disease cases. It illustrates how the dynamics of the disease changed between Phase I and II. Each cluster consists of a 14-weeks window with a circular geographic base which is centered around one of the possible centroids with high RR. For instance, during Phase I, the most-likely cluster of COVID-19 infections was centered at Mexico City metropolitan area with *RR*=4.49, meaning that the population within the cluster was 4.49 times more likely to be exposed to the virus than the population outside the cluster. Interestingly, during Phase II, the most-likely cluster was also centered at Mexico City metropolitan area but with a considerably lower *RR* (*RR*=1.96). Phase II was also characterized by the spread of new infection and death clusters along the country. The findings suggest that during Phase I, both deaths and infections from COVID-19 were concentrated in the country’s capital, however, after the lifting of containment measures, the virus had a rapid spread to the rest of the country due to multiple causes which we further try to investigate.

With respect to the ZINB regression results, we found that in both phases, income inequality, the proportion of adults with obesity and diabetes resulted strongly positively correlated with cumulative COVID-19 infections and deaths. Likewise, annual PM 2.5 concentration (log) was found to be strongly positively correlated to total deaths during Phases I and II. This is, regardless the containment measures, municipalities with deep socio-economic inequalities and poor health outcomes were more likely to suffer from larger infections and deaths during the first wave of the pandemic.

After the introduction, Section II presents our methodological framework. Section III shows the main results and discussion of the empirical findings. Finally, Section IV concludes with some policy recommendations and further work.

## 2. Data and Methods

### 2.1 Spatial scanning statistical analysis

We used Kulldorff’s (1997, 2011) SaTScan analysis to detect space-time clusters of COVID-19 cumulative positive cases and deaths for each of the two phases of lockdown. Under this approach, we tested whether *RR* of the geographical clustering was caused by a random variation or not. This is, whether the expected number of infections (resp. death) within clusters were larger (e.g., cluster of a high risk of infection (resp. death)) than infections (resp. deaths) outside. To do so, we implemented discrete Poisson probability models using different windows of geographical radius. We refer interested readers to Kihal-Talantikite et al. (2013), and Rao et al. (2017) for full technical details. Recent SaTScan applications in the context of COVID-19 can be found in the works of Desjardins et al. (2020) or Kim and Castro (2020) for the cases of U.S. and South Korea, respectively. The most-likely geographical cluster is the one with maximum likelihood, and other clusters with significant log-likelihood ratios (*LLR*) can be seeing secondary potential clusters. Each *LLR* has a *p-value* estimated via 999 Monte Carlo Simulations. Finally, if *p-value<*0.05, then the geographical cluster has significantly higher risk of COVID-19 infections (resp. deaths) than that outside the area.

### 2.2 Potential factors influencing COVID-19 positive cases and deaths

The group of potential predictors is as follows.

- *Socio-economic*. A dummy of metropolitan area, log of population density, proportion of males, Gini of income inequality, and proportion of poverty.
- *Health*. Proportion of obesity in adults aged 20 years or older, proportion of hypertension, proportion of diabetes, and fast food outlets per 1,000 people.
- *Climate*. Log of average monthly maximum temperature, log of average monthly minimum temperature, log of cumulative monthly rainfall, and log of annual PM 2.5 concentration.
- *Commute*. Public buses per 1,000 people, private vehicles automobiles per 1,000 people, and proportion of economic units that are essential activities.

Figure 1 displays the Pearson correlation coefficient resulting from monthly new positive COVID-19 cases and deaths. By visual inspection we notice that population density (log), proportion of adults aged 20 years or older with obesity, annual PM 2.5 (log) and cars per 1,000 people strongly correlate with new monthly COVID-19 cases and monthly deaths.

**Figure 1.**
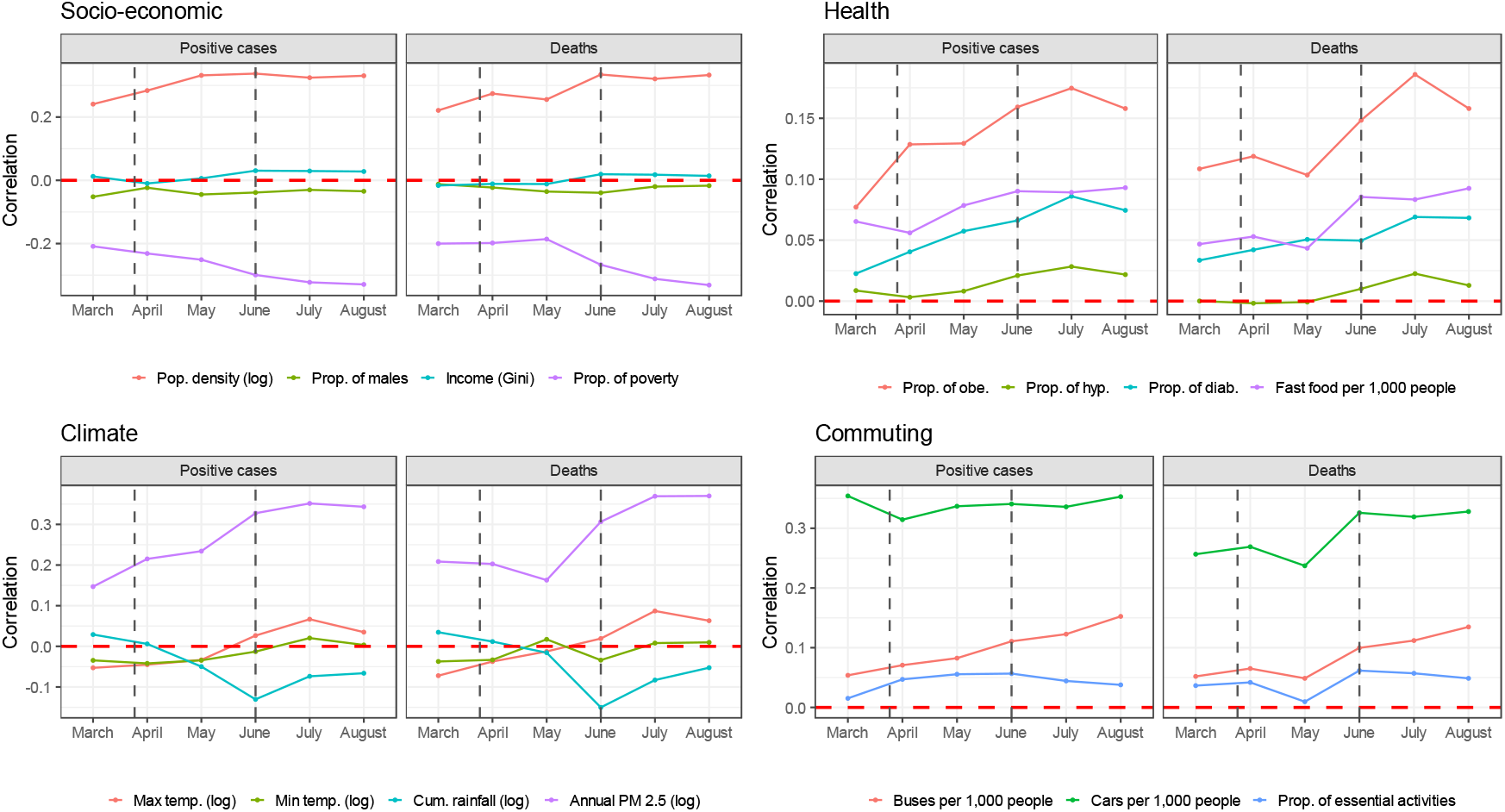
Evolution of Pearson’s correlation between cumulative monthly new positive COVID-19 cases and new deaths using data from 2,459 Mexican municipalities. Vertical dashed lines in black represent the start and end of Phase I.

### 2.3 Econometric strategy

Our econometric strategy is like the one presented in McLaren (2020) in which the dependent variable, *y*, is the cumulative monthly number of reported COVID-19 cases (resp. deaths). More formally, we have:

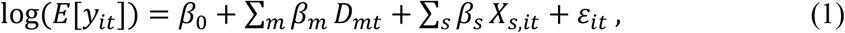

where *y* = 0,1,2 …; *i* stands for municipality and *t* for month. *D*_*mt*_ is a dummy variable that takes the value of 1 if *m =t* and 0 otherwise. *β*_*s*_ are the coefficients associated to the *s* explanatory variables which can be classified into one of the corresponding socio-economic, health, climate or commuting factors.

Our unit of analysis are municipalities, and both the count of positive cases and deaths have more zero observations than predicted by a Poisson process, which assumes that its variance is equal to its mean. For both of our dependent variables the Vuong test detected significant overdispersion, strongly favoring a ZINB regression. The ZINB regression uses the mixing of a process (logistic regression in this study) that predicts some zero counts and another one (negative binomial) that models counts. The logistic analysis of the ZINB model predicts the probability of *y* falling into the zero group (e.g., no positive cases (resp. no deaths) in municipality *i*), while the negative binomial part models the expectation of *y* conditional on a non-zero value. This is, the expected count of the number of positive cases (resp. deaths) in municipality *i*.

## 3. Results

### 3.1 Location of the pandemic in Mexico during Phase I and II

Figure 2(a) depicts the geographical location and size of the pandemic during Phase I. At the beginning of the outbreak, the most-likely cluster of COVID-19 cumulative positive cases was located at Mexico City metropolitan area while secondary clusters were also situated at the center of the country with spatial overlaps.

**Figure 2.**
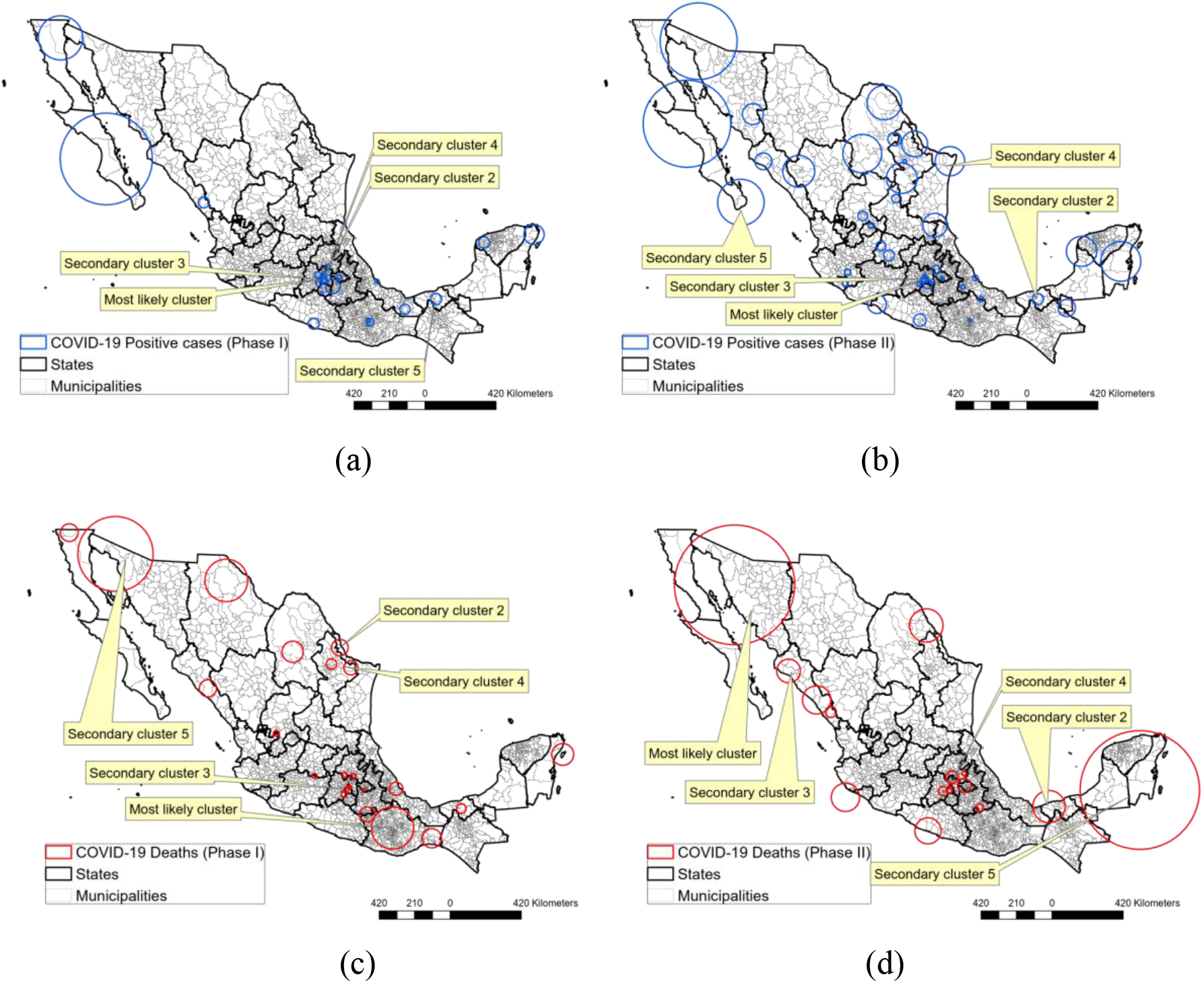
Spatial location of COVID-19 high-risk areas; Cluster of positive cases during (a) Phase I and (b) Phase II; Clusters of deaths during (c) Phase I and (d) Phase II.

More precisely, the most-likely cluster was composed of 21 municipalities (see Table 2) accounting for 29,784 positive cases in a radius of 23.24km. Recall that Mexico City metropolitan area has a total population of more than 20 million inhabitants. Like New York in the U.S., this high-density area was the first in being hit by the pandemic. During this period, the risk of COVID-19 positive cases of the most-likely cluster was about 4.5 (*RR*=4.49) times higher than that in other clusters.

**Table 1.**
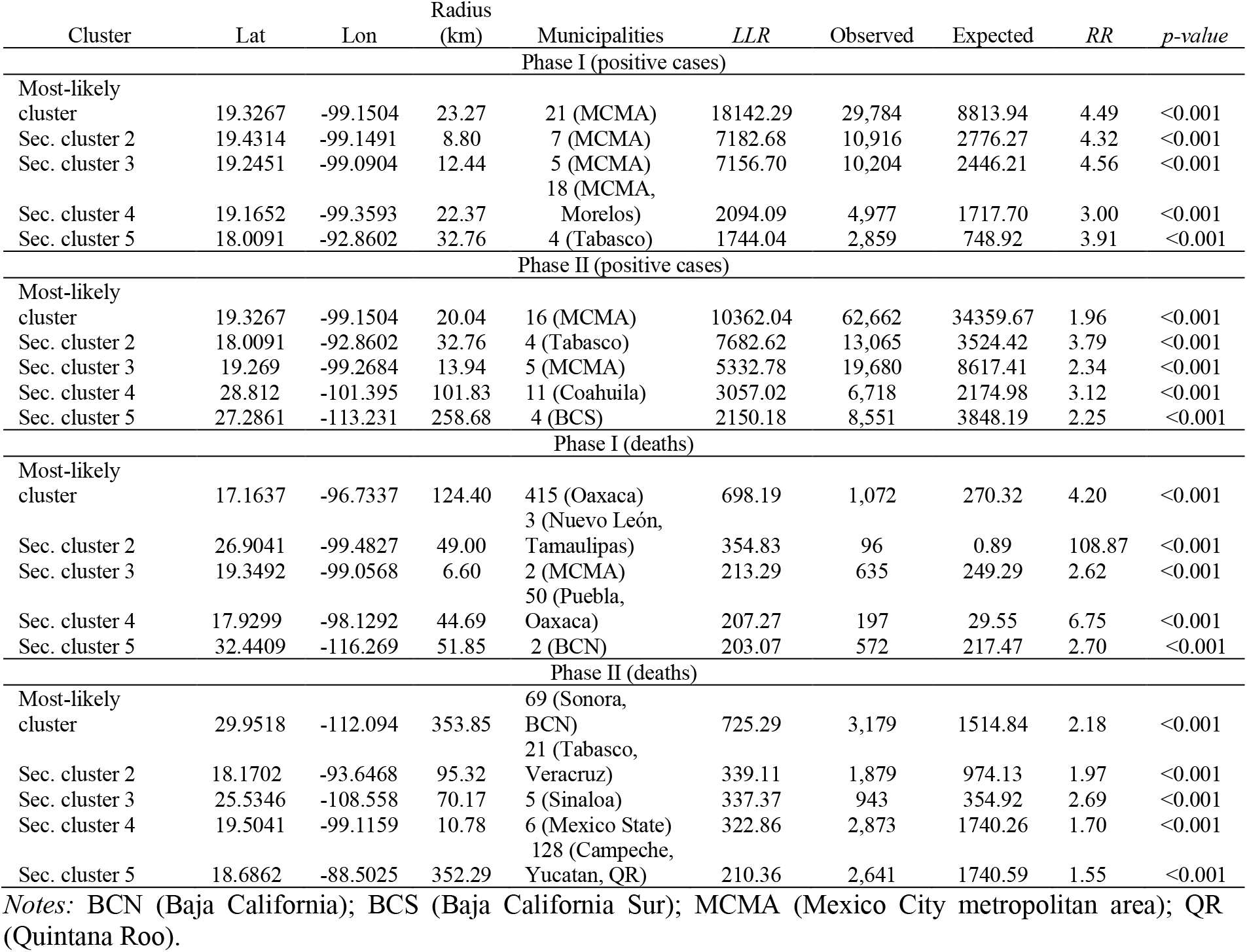
Spatial COVID-19 clusters detected by SaTScan

**Table 2.**
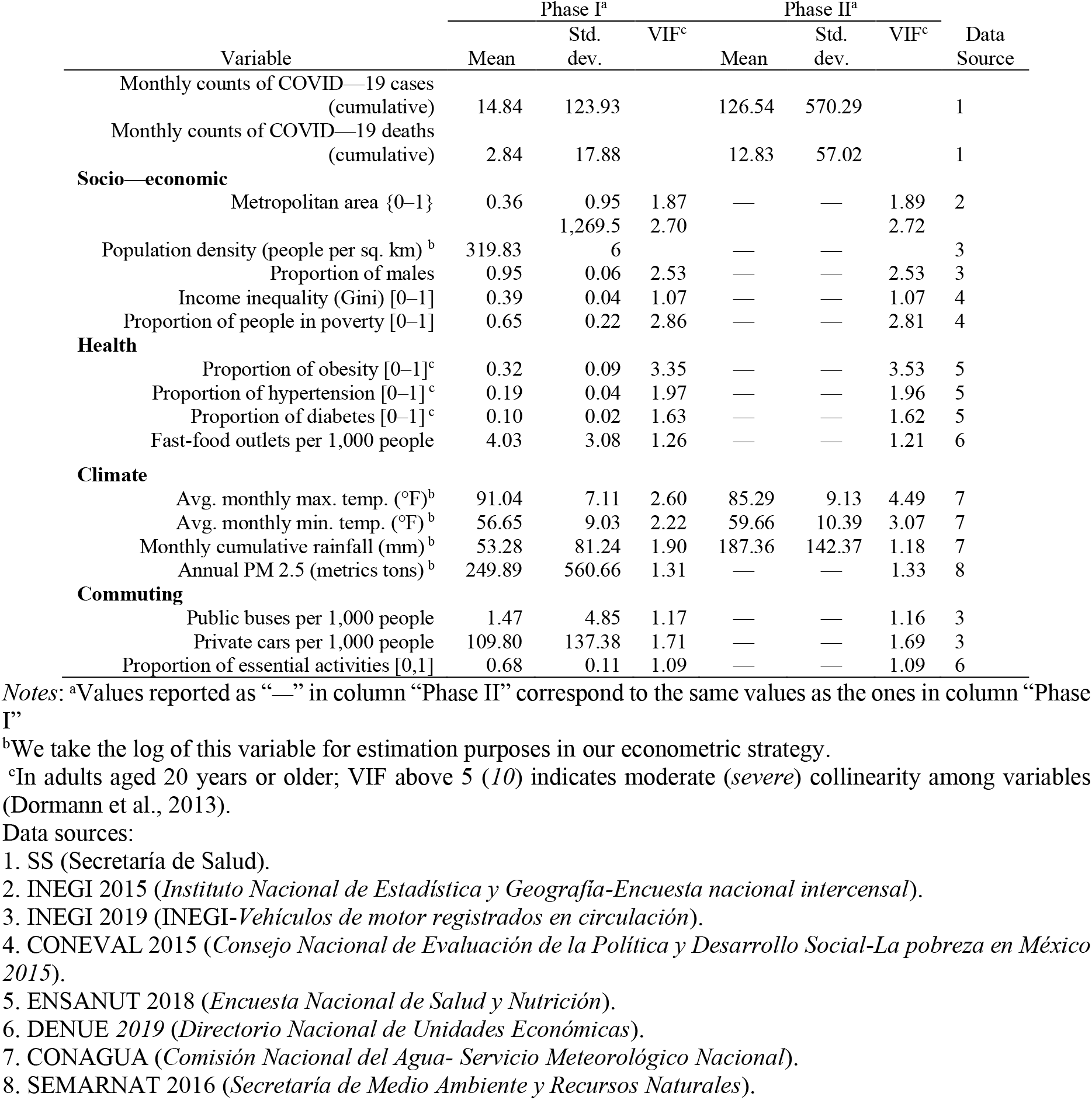
Summary statistics

The effectiveness of social distancing measures imposed by local and federal governments could, perhaps, be appreciated in Figure 2(b). From the figure, we observe that most-likely and secondary clusters shifted towards southern and eastern regions. As expected, these dynamics indicate the rising spread of the virus in less populated areas of the States of Tlaxcala or Guanajuato. Interestingly, however, are the results of spatial clusters of COVID-19 deaths which show an opposite pattern. This is, municipalities clustered at high-risk areas during Phase I were located along different latitudes and longitudes (States of Baja California, Nuevo León, Tamaulipas, Oaxaca and Puebla) without a visible spatial pattern. Alternatively, the high-risk areas during Phase II were concentrated around Mexico City metro area (States of Mexico, Mexico City, Tlaxcala, Puebla, Hidalgo). The center of the most-likely cluster of COVID-19 deaths during Phase II was located in Sonora State, with centroid coordinates 29.9518 latitude and -112.094 longitude (*LLR*=698.19 *p-value<*0.001). This circular area covered 69 municipalities with a radius of 353.85km, and included municipalities sharing border with the U.S. The total number of COVID-19 deaths in the cluster was 3,179, and the risk of fatality was about 2 times (*RR*=2.18) higher than outside this area.

The spatial distribution of COVID-19 positive cases and deaths is likely to be shaped by climate and social processes related to the municipalities’ growth and social inequalities. This motivates our further analysis, which we discuss in the next section regarding to the examination of the main drivers of COVID-19 disease spread.

### 3.2 Descriptive statistics of the variables incorporated in the model

During the Social Distancing Program, a total of 1,493 (60.71%) municipalities reported at least one confirmed case, and 1,216 (50.5%) reported at least one death. Three months after the Social Distancing Program, 2,096 (85.2%) and 1,551 (63%) municipalities reported at least one confirmed case and deaths, respectively.

Table 2 presents the summary statistics of the variables of interest. As we can notice, due to data availability only some climate-related variables present monthly changes in their values. Among other facts, 36 ± 95% (mean ± standard deviation) of the municipalities belong to metropolitan areas, the average Gini income inequality per municipality is 0.39±0.04 while in the average municipality 65±22% of the population lives below the poverty line. In average, Mexican municipalities have 319.83 ± 1,269.56 inhabitants per sq. km of land, but its standard deviation is considerably large indicating that datapoints tend to be far from the mean value.

The proportion of obesity is very high, this is, in average 32 ± 9% of the population aged 20 years or older suffer from obesity. As mentioned previously, the relationship between obesity and respiratory disease is largely known (Walton et al., 2009; Jiao et al., 2015; Richardson et al., 2015) and we expect a positive relationship with COVID-19 deaths. However, the magnitude of the coefficient estimates associated to obesity, as well as the coefficients of the other comorbidities, is region dependent.

In the category of environmental variables, we have converted temperature into Fahrenheit degrees to avoid negative values of Celsius degrees during cold months so that the log transformation does not disrupts our analysis. Long-term exposure to PM 2.5 adversely affects respiratory and cardiovascular systems and may increases lethality rates of COVID-19 (Wu et al., 2020). This could be particularly true for municipalities in metro areas. In Méndez-Arriaga (2020) it has been documented that climate conditions in States with low temperatures and poor precipitation (in mm), such as Tlaxcala, Guanajuato or Michoacán, favoured local infections during Phase I.

Our proxies for human mobility behaviour are expected to be positively correlated to the spread of the virus (see Figure 1), but its association with deaths remains unknown. The variable proportion of essential activities include health-related companies, public security and fundamental sectors of the economy (utilities, food and beverages, agriculture, fisheries and livestock, industrial chemicals, storage, courier services, private security, telecommunications, emergency services, funeral services, transportation of individuals, logistics and governmental activities). The average percentage of economic units that operate essential activities during Phase I was 68 ± 11%.

### 3.3 Main determinants of COVID-19 cumulative cases and deaths during Phase I and II

For the easiness of synthesizing our results, Figure 3 shows the effect size of the estimated coefficients related to the count part of the model, e.g., negative binomial, and Tables 3 and 4 summarize full results. Recall that the count model describes associations between explanatory variables and COVID-19 cases (deaths) among municipalities with at least one reported COVID-19 case (death). During both Phases I and II, we observed (Figure 3(a)) a large and significant positive association between municipal-level cumulative positive cases and proportion of obesity (*p-value<*0.001), proportion of diabetes (*p-value<*0.001) and income inequality (*p-value<*0.001). Although positive and statistical significative, the size of the estimated coefficients of proportion of essential activities, population density (log), average minimum temperature (log) and annual PM 2.5 concentration (log) is considerably smaller.

**Table 3.** Incidence rate ratios (IRR) of cumulative COVID-19 cases

|  |  | Phase I |  | Phase II |  |
| --- | --- | --- | --- | --- | --- |
|  |  | IRR | p-value | IRR | p-value |
| *Count Model* |  |  |  |  |  |
|  | Intercept | 4.92E-06 | <0.001 | 7.54E-4 | <0.001 |
|  | Month 2 | 14.42 | <0.001 | 2.532 | <0.001 |
|  | Month 3 | 111.26 | <0.001 | 3.54 | <0.001 |
| Socio-economic |  |  |  |  |  |
|  | Metropolitan area | 1.11 | 0.266 | 1.23 | <0.001 |
|  | Pop. density (log) | 1.71 | <0.001 | 1.58 | <0.001 |
|  | Prop. of males | 0.46 | 0.451 | 0.01 | <0.001 |
|  | Income inequality (Gini) | 4,045.05 | <0.001 | 10,899.25 | <0.001 |
|  | Prop. of people in poverty | 0.12 | <0.001 | 0.05 | <0.001 |
| Health |  |  |  |  |  |
|  | Prop. of obesity | 658.14 | <0.001 | 327.83 | <0.001 |
|  | Prop. of hypertension | 0.01 | <0.001 | 0.01 | <0.001 |
|  | Prop. of diabetes | 394.17 | <0.001 | 6,932.12 | <0.001 |
|  | Fast-food per 1,000 people | 1.01 | 0.652 | 1.02 | 0.005 |
| Climate |  |  |  |  |  |
|  | Avg. monthly max. temp. (log) | 0.96 | 0.952 | 3.38 | <0.001 |
|  | Avg. monthly min. temp. (log) | 1.2 | 0.606 | 1.25 | 0.102 |
|  | Monthly cumulative rainfall (log) | 1.03 | 0.29 | 0.87 | <0.001 |
|  | Annual PM 2.5 (log) | 1.77 | <0.001 | 1.91 | <0.001 |
| Commuting |  |  |  |  |  |
|  | Public buses per 1,000 people | 1.01 | 0.058 | 1.02 | <0.001 |
|  | Private cars per 1,000 people | 1.01 | <0.001 | 1.01 | <0.001 |
|  | Prop. of essential activities | 1.16 | 0.659 | 0.54 | <0.001 |
| *Zero-Inflated Model* |  |  |  |  |  |
|  | Intercept | 2.44E-06 | 0.17 | 0.04 | 0.806 |
|  | Month 2 | 0.04 | <0.001 | 0.41 | 0.045 |
|  | Month 3 | 0.01 | <0.001 | 0.27 | 0.004 |
| Socio-economic |  |  |  |  |  |
|  | Metropolitan area | 0.12 | 0.002 | 1.68E-05 | 0.922 |
|  | Pop. density (log) | 0.89 | 0.327 | 1.46 | 0.014 |
|  | Prop. of males | 3.07E+09 | <0.001 | 0.48 | 0.826 |
|  | Income inequality (Gini) | 0.08 | 0.655 | 7.17E-05 | 0.016 |
|  | Prop. of people in poverty | 129.78 | 0.13 | 8.51E+04 | <0.001 |
| Health |  |  |  |  |  |
|  | Prop. of obesity | 0.03 | 0.322 | 1.57 | 0.886 |
|  | Prop. of hypertension | 1289.48 | 0.05 | 3.61E+15 | <0.001 |
|  | Prop. of diabetes | 0.07 | 0.748 | 1.98E-17 | <0.001 |
|  | Fast-food per 1,000 people | 1.06 | 0.282 | 1.07 | 0.082 |
| Climate |  |  |  |  |  |
|  | Avg. monthly max. temp. (log) | 0.11 | 0.59 | 0.08 | 0.499 |
|  | Avg. monthly min. temp. (log) | 1.61 | 0.808 | 0.87 | 0.923 |
|  | Monthly cumulative rainfall (log) | 1.06 | 0.587 | 1.39 | 0.185 |
|  | Annual PM 2.5 (log) | 0.6 | 0.001 | 0.54 | 0.001 |
| Commuting |  |  |  |  |  |
|  | Public buses per 1,000 people | 1.01 | 0.824 | 0.01 | 0.001 |
|  | Private cars per 1,000 people | 1.01 | 0.004 | 0.94 | <0.01 |
|  | Prop. of essential activities | 40.81 | 0.164 | 0.59 | 0.521 |
| Observations |  | 7,390 |  | 7,390 |  |

**Table 4.**
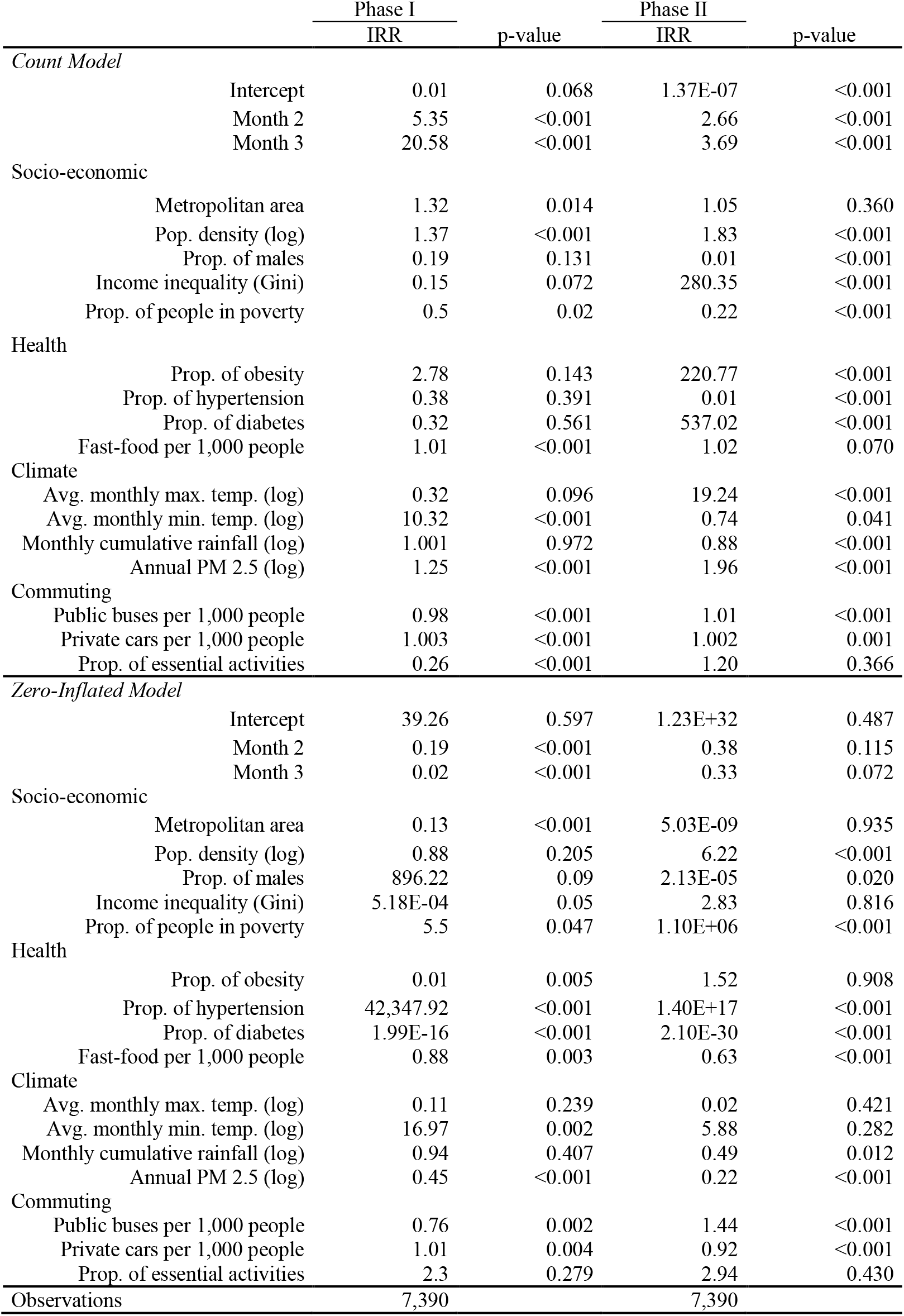
Incidence rate ratios (IRR) of COVID-19 deaths

|  |  | Phase I |  | Phase II |  |
| --- | --- | --- | --- | --- | --- |
|  |  | IRR | p-value | IRR | p-value |
| Count Model |  |  |  |  |  |
|  | Intercept | 0.01 | 0.068 | 1.37E-07 | <0.001 |
|  | Month 2 | 5.35 | <0.001 | 2.66 | <0.001 |
|  | Month 3 | 20.58 | <0.001 | 3.69 | <0.001 |
| Socio-economic |  |  |  |  |  |
|  | Metropolitan area | 1.32 | 0.014 | 1.05 | 0.360 |
|  | Pop. density (log) | 1.37 | <0.001 | 1.83 | <0.001 |
|  | Prop. of males | 0.19 | 0.131 | 0.01 | <0.001 |
|  | Income inequality (Gini) | 0.15 | 0.072 | 280.35 | <0.001 |
|  | Prop. of people in poverty | 0.5 | 0.02 | 0.22 | <0.001 |
| Health |  |  |  |  |  |
|  | Prop. of obesity | 2.78 | 0.143 | 220.77 | <0.001 |
|  | Prop. of hypertension | 0.38 | 0.391 | 0.01 | <0.001 |
|  | Prop. of diabetes | 0.32 | 0.561 | 537.02 | <0.001 |
|  | Fast-food per 1,000 people | 1.01 | <0.001 | 1.02 | 0.070 |
| Climate |  |  |  |  |  |
|  | Avg. monthly max. temp. (log) | 0.32 | 0.096 | 19.24 | <0.001 |
|  | Avg. monthly min. temp. (log) | 10.32 | <0.001 | 0.74 | 0.041 |
|  | Monthly cumulative rainfall (log) | 1.001 | 0.972 | 0.88 | <0.001 |
|  | Annual PM 2.5 (log) | 1.25 | <0.001 | 1.96 | <0.001 |
| Commuting |  |  |  |  |  |
|  | Public buses per 1,000 people | 0.98 | <0.001 | 1.01 | <0.001 |
|  | Private cars per 1,000 people | 1.003 | <0.001 | 1.002 | 0.001 |
|  | Prop. of essential activities | 0.26 | <0.001 | 1.20 | 0.366 |
| Zero-Inflated Model |  |  |  |  |  |
|  | Intercept | 39.26 | 0.597 | 1.23E+32 | 0.487 |
|  | Month 2 | 0.19 | <0.001 | 0.38 | 0.115 |
|  | Month 3 | 0.02 | <0.001 | 0.33 | 0.072 |
| Socio-economic |  |  |  |  |  |
|  | Metropolitan area | 0.13 | <0.001 | 5.03E-09 | 0.935 |
|  | Pop. density (log) | 0.88 | 0.205 | 6.22 | <0.001 |
|  | Prop. of males | 896.22 | 0.09 | 2.13E-05 | 0.020 |
|  | Income inequality (Gini) | 5.18E-04 | 0.05 | 2.83 | 0.816 |
|  | Prop. of people in poverty | 5.5 | 0.047 | 1.10E+06 | <0.001 |
| Health |  |  |  |  |  |
|  | Prop. of obesity | 0.01 | 0.005 | 1.52 | 0.908 |
|  | Prop. of hypertension | 42,347.92 | <0.001 | 1.40E+17 | <0.001 |
|  | Prop. of diabetes | 1.99E-16 | <0.001 | 2.10E-30 | <0.001 |
|  | Fast-food per 1,000 people | 0.88 | 0.003 | 0.63 | <0.001 |
| Climate |  |  |  |  |  |
|  | Avg. monthly max. temp. (log) | 0.11 | 0.239 | 0.02 | 0.421 |
|  | Avg. monthly min. temp. (log) | 16.97 | 0.002 | 5.88 | 0.282 |
|  | Monthly cumulative rainfall (log) | 0.94 | 0.407 | 0.49 | 0.012 |
|  | Annual PM 2.5 (log) | 0.45 | <0.001 | 0.22 | <0.001 |
| Commuting |  |  |  |  |  |
|  | Public buses per 1,000 people | 0.76 | 0.002 | 1.44 | <0.001 |
|  | Private cars per 1,000 people | 1.01 | 0.004 | 0.92 | <0.001 |
|  | Prop. of essential activities | 2.3 | 0.279 | 2.94 | 0.430 |
| Observations |  | 7,390 |  | 7,390 |  |

**Figure 3.**
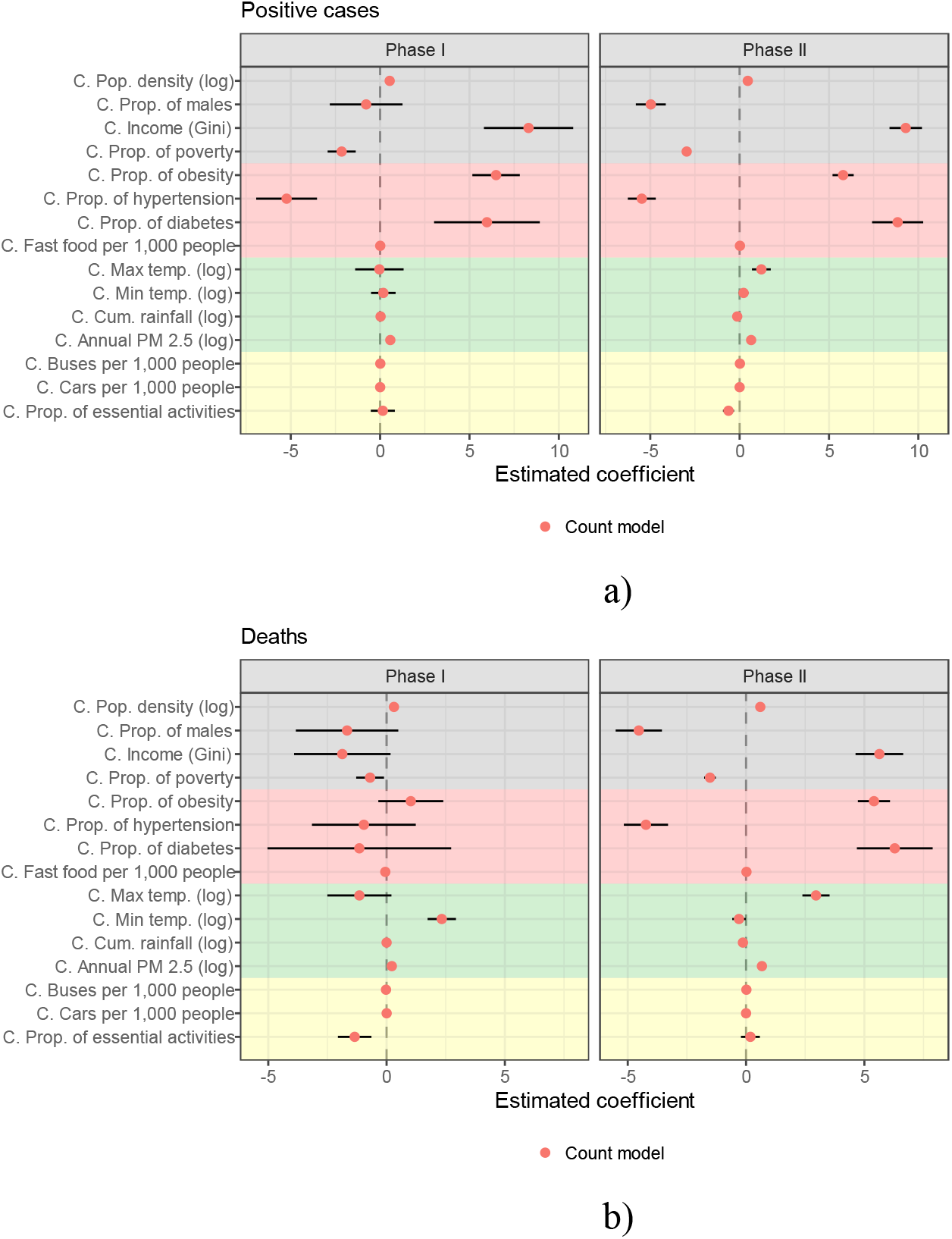
Effect size of predictor coefficients, count model of the Zero-Inflated Negative Binomial. Black lines represent the 95% confidence interval. Variables in grey, red, green and yellow color correspond to socio-economic, health, climate and commuting factors, respectively

In regard to COVID-19 fatalities, population density (log) and average minimum temperature (log) were found to be positively associated to deaths. The statistical significance of population density is not surprising as we could hypothesized such result from the strong positive correlation shown in Figure 1. However, positive association between average minimum temperature and COVID-19 deaths is less obvious. This illustrates the practicality of ZINB models. Socio-economic and health-related variables had an important role on the evolution of monthly cumulative number of COVID-19 cases and deaths. For the count model results in Figure 3, the estimated coefficient of population density (log) during Phase I equals 0.53 (resp. 0.31) and it is statistically significant at the 99%. An alternative explanation for the estimated coefficients is the incidence rate ratios (*IRR*), also known as odds ratios in ZINB models, which are calculated by taking the exponent of the estimated coefficients in Figure 3. The *IRR* of population density (log), see Table 3, indicates that a unit increase in this variable increased it by 1.71 (=exp(0.53)) (resp. 1.37) times among those municipalities who experienced cases of coronavirus disease 2019 infection (resp. deaths) during the Phase I. In Phase II, however, the magnitude of the estimated coefficient became larger in the case of deaths, meaning that more densely populated municipalities suffered from larger counts of fatalities. This result is also partially illustrated in Figure 2. The finding is in line with recent published work using U.S. county-level data (Sun et al., 2020a) or Iran province-level data (Ahmadi et al. 2020) but contrary to the empirical evidence of the provincial regions in China (Sun et al., 2020c).

Municipalities located at metropolitan areas were more-likely (*p-value<*0.001) to suffer from infections and deaths in Phase II. Income inequality is revealed as one of main factors associated with the spread of the virus and deaths. During Phase I, holding all other municipal-level variables at their mean values, positive cases increased by a factor of 4,045.05 for each one unit increase in the Gini index, while in Phase II this factor increased to 10,178.66. Moreover, municipalities with disproportionately social inequalities (e.g., municipalities in the States of Oaxaca, Mexico or Mexico City) suffered from larger *IRR* of COVID-19 deaths. The result is in line with the evidence of the U.S. at the county-level (Millett, 2020) or Singapore at the planning area-level (Yi et a., 2020). The pronounced social disparities across states has shown that vulnerable geographical areas in terms of large social disadvantages were those with the highest numbers of cases and deaths of COVID-19.

Our findings also provide support that poor air quality conditions at the municipal-level strongly correlate to the prevalence and deaths due to COVID-19. During Phase I (resp. Phase II) an increase in 1 metric tonne in PM2.5 (log) per year would increase COVID-19 death rate by a factor of 1.25 (resp. 1.97). The finding is in line with relevant studies tackling the atmospheric determinants of population’s vulnerability to COVID-19 in the U.S. (Bashir et al., 2020; Wu et al., 2020), Italy (Lubrano et al., 2020), China (Lin et al., 2020a) or some cities of Latin America (Bolaño et al., 2020), all of them unanimously confirmed the correlation between atmospheric conditions and COVID-19 cases, making poor air quality an additional co-determinant of COVID-19 lethality. With respect to precipitation, we observed that high cumulative rainfall (log) reduced the virus transmission and deaths during Phase II, only. Different from air pollution, the literature focusing on the role of amount of rainfall and virus transmission/deaths have found inconclusive results. In some countries like the U.S. (Bashir et al., 2020), Indonesia (Tosepu et al., 2020) or Brazil (Rosario et al., 2020) rainfall has been shown to be not associated with the dynamics of the virus.

The large magnitude of the coefficients associated to health-related explanatory variables during Phase I indicates that regions with high prevalence of comorbidities, such as municipalities in the states of Mexico, Sinaloa, Veracruz or Yucatan, were hard hit by COVID-19. Note that during Phase I comorbidities where not statistically associated to the cumulative count of deaths.

Lastly, commuting-related factors are found to be weakly correlated to confirmed cases and deaths. For instance, during Phase I, municipalities with large proportion of essential activities reported lower death rates (*IRR*= 0.26, *p-value<*0.001). After the social distancing was gradually lifted in Phase II, an increase in the proportion of essential activities was linked to a decrease (*IRR*= 0.46, *p-value<*0.001) in the spread of the virus. The explanation for these apparently inconsistent effects is tied to the share of the informal employment in Mexico (53.4% in 2016, according to the International Labour Organization (ILO, 2018)). Needless to say that the size of the informal employment in the country is comparable to other Latin American nations (Argentina, 47.2%; Brazil 40.5%; Chile 40.1%; Ecuador 59%) or China (54.4%).

## 3. Concluding remarks

### 4.1 Discussion

The spatial distribution of emerging space-time clusters of COVID-19 at the municipal-level showed that after Phase I was lifted, the de-escalation process exhibited rapidly spread of the virus to northern and southern regions, particularly to those municipalities along the Mexico-U.S. border. We identified a total of 23 clusters of COVID-19 infections during Phase I, and surprisingly, the total number of clusters increased to 40 in Phase II. Notwithstanding, we also found an opposite dynamic with respect to cumulative COVID-19 deaths. This is, the total number of clusters reduced from 24 to 16 between Phase I and II, respectively, and their centroids moved away from Mexico City metropolitan area to Sonora and other bordering states. Although Phase II was characterized by a small number of clusters of COVID-19 deaths, they also reported a considerably larger number of cumulative deaths. For instance, the most-likely cluster during Phase I reported 1,072 deaths whereas in Phase II, it exhibited 3,179. Our spatial analysis illustrates the usefulness of the spatial scan statistic (SaTScan) as a tool for geographical disease surveillance.

Aside from the physical restriction measures imposed by local and federal governments, other potential municipal-level factors associated to dynamic of the disease could be existing socio-economic inequalities. The empirical evidence of Das et al. (2020) showed that COVID-19 spread grow exponentially in those Indian regions with large socio-economic deprivation. Similarly, the evidence comparing United Kingdom regions (Sun et al., 2020b) revealed important geographical disparities in risk and outcomes of the disease. This is, the risk of dying among those diagnosed with the infection was higher for those belonging to minority ethnic groups and those living in more deprived areas. Sannigrahi et al. (2020) also found strong evidence supporting the idea that spatial determinants of COVID-19 infections and deaths in Europe were mainly associated to poverty rates and income inequality.

Additional arguments explaining the geographical spread of the disease are presented in Geng et al. (2020). Their study found associations between reopening of urban business centers in the U.S. and rapid spread of the virus across counties. Sun et al. (2020a) highlighted the noticeable spatial heterogeneity in the distribution of positive cases in the U.S., in particular, when comparing counties in the center of the country with those located at the East and West coast. The authors illustrated the strong positive correlation between socio-economic factors (population density, social segregation, and differences by ethnicity) and infection rate. The link between space-time variations of disease and socio-economic factors has also recently been investigated by Snyder and Parks (2020), Xie et al. (2020) and Karaye and Horney (2020).

In the context of Mexico, our results at the municipal-level indicate that, regardless of lockdown policies, metabolic disorders, such as obesity or diabetes, are able to exacerbate both infections and deaths. Our findings are in line with those presented in Lubrano et al. (2020), in which the extremely high level of COVID-19 lethality in Northern Italy compared to the rest of the country can be explained by differences in air pollution and obesity. The above-mentioned link, between obesity/air pollution and higher COVID-19 lethality is also supported by Sanchis-Gomar et al. (2020). The authors detail how adipose tissue may be vulnerable to COVID-19 infection, therefore, obese patients also have worse outcomes with COVID-19 infection, including respiratory failure, need for mechanical ventilation, and higher mortality (Almerie and Kerrigan, 2020; Palaiodimos et al., 2020; Flint and Tahrani, 2020). Similarly, the presence of diabetes has been associated with an increased mortality risk (Pitocco et al., 2020; Papadokostaki et al., 2020) because people with diabetes could suffer from a loss of capacity to regulate immunity. Therefore, due to the large prevalence of diabetes (15%) and obesity (60%) in Mexico, we expect COVID-19 lethality being considerably higher than the one experienced by other countries in the region. In this vein, our variable fast food per 1,000 people was intended to discover possible effects of fast food restaurants on infections and deaths. There is an ongoing debate on weather higher density of unhealthy food options is associated with higher obesity and diabetes prevalence. While the evidence is mixed (Frankenfeld et al., 2015, Mejova et al., 2015), Mexico has recently implemented a front-of-pack labeling system as an attempt to tackle obesity by reducing the purchase of unhealthy products. All in all, our empirical evidence does not support the idea that an increasing number of local fast food restaurants is associated with higher rates of disease infections or lethality.

### 4.2 Policy recommendations

On the one hand, the detection of space-time clusters of infections and deaths, Figure 2, can be exploited by policymakers and healthcare authorities to target regions that are on need of priority interventions. For instance, when there is a substantial overlap of secondary clusters, e.g., Figure 2(a), it is indicative of an urgent need of regional measures for the prevention, containment and coordination to be implemented in combating the COVID-19 health crisis. Successfully examples of localized policy interventions are detailed in Leal-Neto et al. (2020) and Dlamini et al. (2020) for the case of Brazil and Eswatini (Africa), respectively. Likewise, some of the spatial clusters identified in this work grouped together municipalities from different states. This scenario is of key relevance while designing localized public health efforts. In Mexico, the federal government has delegated to Federal States much of the responsibility about the management of the public health system during the pandemic. Federal States also have main duties and responsibilities related to the restriction of non-essential economic activities when the Epidemiological Risk Traffic Light (a bi-weekly monitoring system met to monitor and grade the use of public space according to virus transmission) turns into red color. In context of emerging clusters sharing municipalities from different states, coordinated efforts among various governments levels are critical. Past published research has shown the advantages of these type of coordinated public policy interventions not only to prevent the spread of sexual transmission diseases (Marotta, 2016) or Salmonella (Seixas et al., 2018) but also to provide with better healthcare services to vulnerable population suffering from cleft lip and palate (Gasca et al., 2019), obesity (Gartner et al., 2016), tuberculosis (Rao et al., 2017), and so forth.

On the other hand, while some experts say that the hardest hit countries had an aging population (Gardner et al., 2020), underdeveloped healthcare systems (Tanne et al., 2020) or unfavorable natural environment (Di Marco et al., 2020), it seems that unresolved structural problems of high poverty and income inequality aggravated the spread of the pandemic in Mexico. Thus, government response policies, such as testing procedures, tracking of individuals, social distancing measures or plans for hospital reconversion and immediate expansion will not be sufficient to mitigate the dynamics of the disease spread. Unless major policy changes are implemented to improve environmental sustainability and promote human development, more disadvantaged Mexican municipalities will become hotspots for virus circulation and re-emergence. More precisely, some of the possible policy actions could include support for workers in the informal sector, assistance for the population in poverty or a decarbonization pathway that focuses on electrification and clean renewable energy. Interestingly, the recent front-of-pack labeling system based on warning labels of unhealthy products seems to be an adequate government’s strategy to fight obesity and ultimately COVID-19.

## Data Availability

All data are drawn from public databases. References to these databases are cited in the text and listed in the bibliography.

## Credit Author Statement

Francisco Benita: Conceptualization, Methodology & Writing. Francisco Gasca-Sanchez: Conceptualization, Data curation, Visualizations.

